## supplementary information for "Risk averse reproduction numbers improve resurgence detection"

### S1 Appendix

##### Real-time estimates of reproduction numbers

The estimates of transmissibility underpinning **Figs 3-6** of the main text use the maximally informed posterior distributions $\mathbf{P}(R_j(t)|I_1^T)$, which provide a real-time estimate for $R_j(t)$ at the present, $R_j(T)$, but informs earlier $R_j(t)$ estimates using past and future (i.e., $I_j(s): t \leq s \leq T$) incidence data. This has only a minor effect on their qualitative interpretation about 1 as shown in [1] and allows us to summarise the entire time series of estimates with a single plot. However, to ensure that our claims are representative of what would be made based on real-time estimates we perform additional sensitivity analyses at key time points for the case study of COVID-19 for 20 cities in Israel in **Fig A**.

There we limit $T$ (endpoints of panels) to various times that coincide with the growth of the Delta strain (dot-dashed black) and infer the standard and risk averse reproduction numbers $\hat{X}(t)$ for $X = R, E$, as well as their corresponding probabilities of resurgence $\mathbf{P}(\hat{X}(t) > 1)$. As expected, and in line with the results in **Fig 5** of the main text, we find that the risk averse $E$ would support an earlier initiation of the booster campaign (start time shown by a dashed grey vertical line) than the standard effective $R$. There are also substantial lags until when $R$ signals resurgence ($\mathbf{P}(\hat{X}(t) > 1) > \frac{1}{2}$), at which point the Delta strain has already become firmly established. The larger credible intervals of $E$ for the second panel reflect that several cities had very small incidence. In contrast, $R$ somewhat over-smooths this effect.

##### General risk averse properties of *E*

In the main text we showed that *E* has risk averse properties because it is the solution of an E-optimal design that tends to up-weight groups that are likely to cause resurgences. While the E-optimal property depends on the statistical renewal models we used, here we provide two explanations as to why *E* will maintain its risk averse properties even when applied more generally. First, assume that we hold the $R_j$ of our groups constant and project forwards in time (a standard assumption when forecasting infectious diseases [2,3]). As time passes, we would find that the rank (by magnitude) of the $\Lambda_j$ becomes progressively correlated with the rank of the $R_j$. This means that the group with the largest local reproduction number tends, eventually, to also have the largest circulating infection count.

Consequently, we gain some predictive insight if we can meaningfully assign weights to match the ranks of the $R_j$ while incorporating local estimate uncertainty. *E* does this directly by setting weights to match the fractional rank of $R_j$ and factors in $R_j$ estimate uncertainties within its weighted formulation. In reality, the $R_j$ will change with time but *E* will also update to reflect these variations. Hence, *E* provides some predictive insight into what can happen given the current state of local groups and encodes a risk averse property since statistically riskier groups (i.e., groups with larger $R_j$ that have higher chances of rapid spread and lower chances of infections naturally fading out) will dominate those forecasts.

Second, *E* is the contraharmonic mean of the $R_j$. This is equally a Lehmer mean with power $q = 2$. Lehmer means are defined as $\left(\sum_{j=1}^{p} R_j^q\right)\left(\sum_{j=1}^{p} R_j^{q-1}\right)^{-1}$[4] and commonly applied in signal processing as envelope detectors (i.e., they highlight peaks in waveforms). Further, these means interpolate between the arithmetic mean ($q = 1$) and the maximum of their inputs ($q \to \infty$), which respectively relate to *D* and the max $R_j$ metrics that we explored in the main text. Accordingly, *E* will always have a risk averse property without being overly sensitive like means with larger $q$, which include the max $R_j$ statistic. In contrast, *R*, which is a weighted form of the $q = 1$ mean has no guaranteed resurgence detection properties.

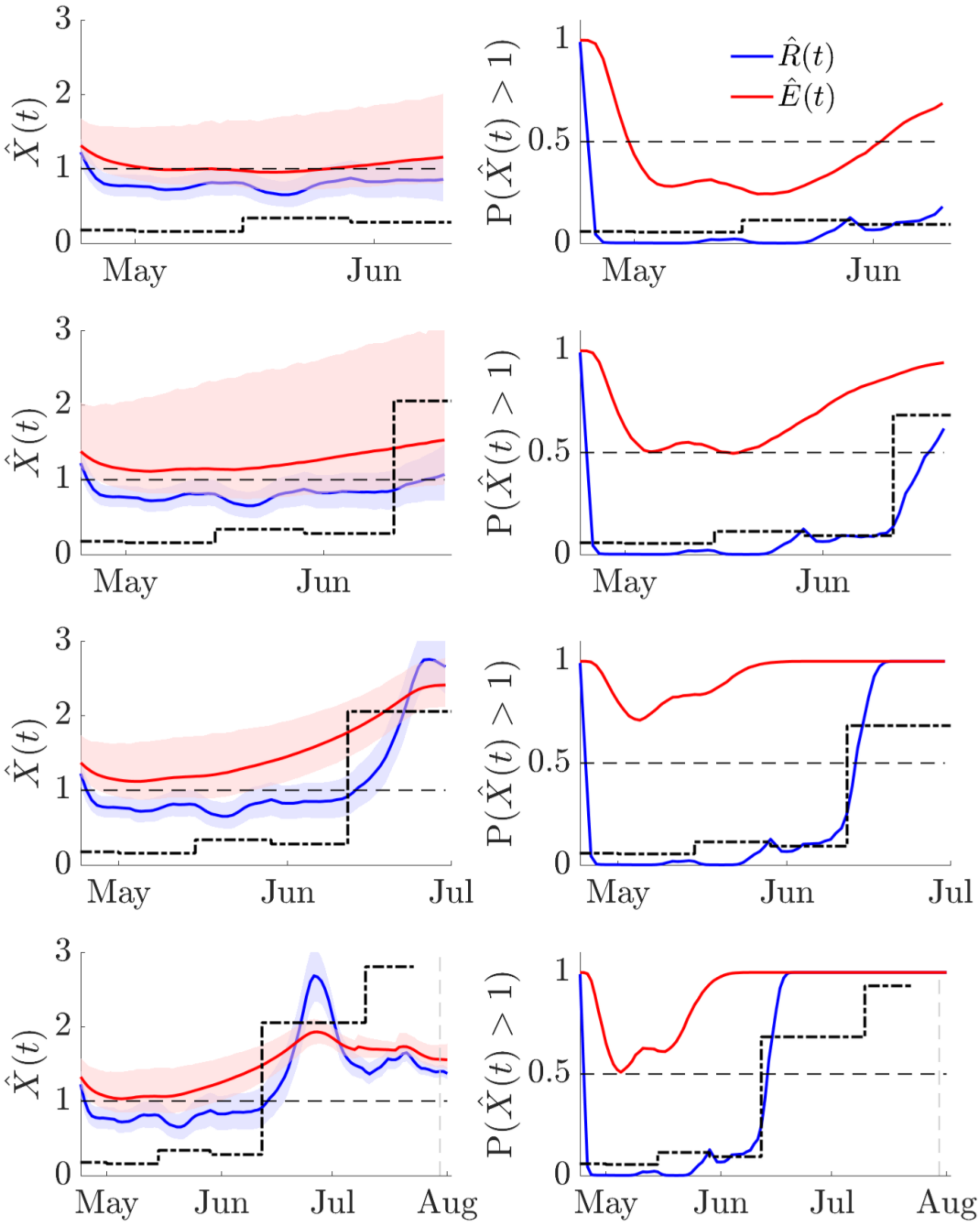


**Fig A: Real-time analysis of COVID-19 Delta strain dynamics in Israel**. We repeat the analysis of **Fig 5** of the main text but truncate our analysis to key points in the time series to provide real-time or prospective estimates of transmissibility. Left panels provide mean estimates $\hat{X}(t)$ and their 95% credible intervals for standard (red, $R$) and risk averse (blue, $E$) reproduction numbers. The dot-deashed black line indicates the proportion of Delta strain cases (values are between 0-1, these are shown on right panels but scaled up on left panels for comparison) and the grey dashed line shows when the booster campaign began. Right panels plot corresponding resurgence probabilities $\boldsymbol{P}(\hat{X}(t) > 1)$.

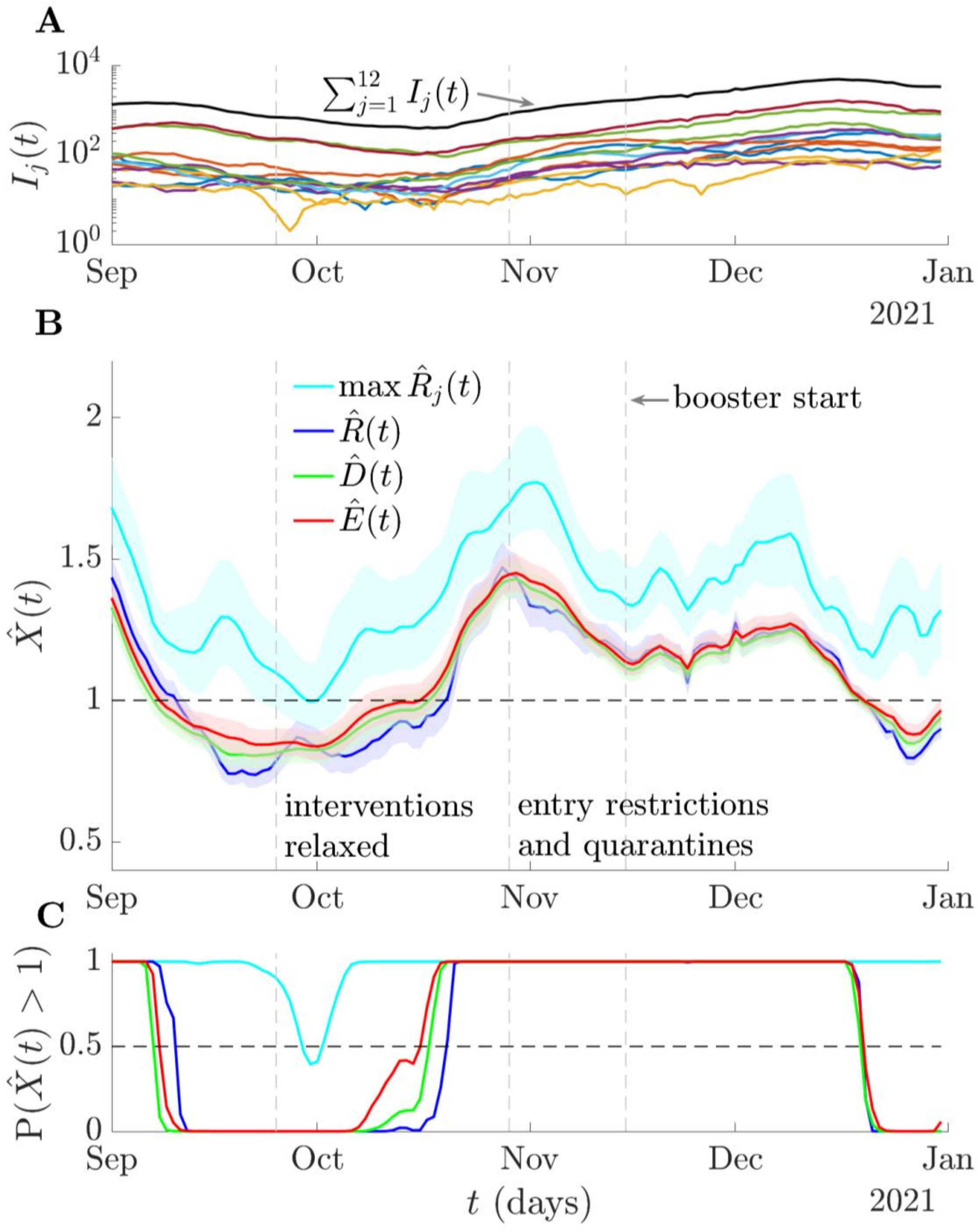


**Fig B: Risk averse reproduction numbers for COVID-19 in Norway.** We plot cases by date of positive test (and in log scale) in (A) for $p = 12$ districts in Norway for COVID-19 in 2021 from [5] with summed incidence in black. We also plot these curves in **Fig C** below. As in **Fig 5** of the main text, we infer standard, $\hat{R}(t)$, maximum group, max $\hat{R}_j(t)$, mean, $\widehat{D}(t)$, and risk averse, $\hat{E}(t)$, reproduction numbers (with 95% credible intervals) using EpiFilter [6] in (B) with the serial interval distribution estimated in [7]. We plot intervention relaxation and implementation times from [5] as vertical dashed lines. We assume stable reporting and that generation times are well approximated by serial intervals. (C) integrates posterior estimates from (B) into resurgence probabilities $\boldsymbol{P}(\hat{X}(t) > 1)$. All the reproduction numbers evidence a need for intervention imposition earlier than November but $\hat{E}(t)$ signals this need notably earlier than $\hat{R}(t)$. The max $\hat{R}_j(t)$ estimate is not informative. All other estimates support the intervention relaxation time and emphasise the need for sustained boosters.

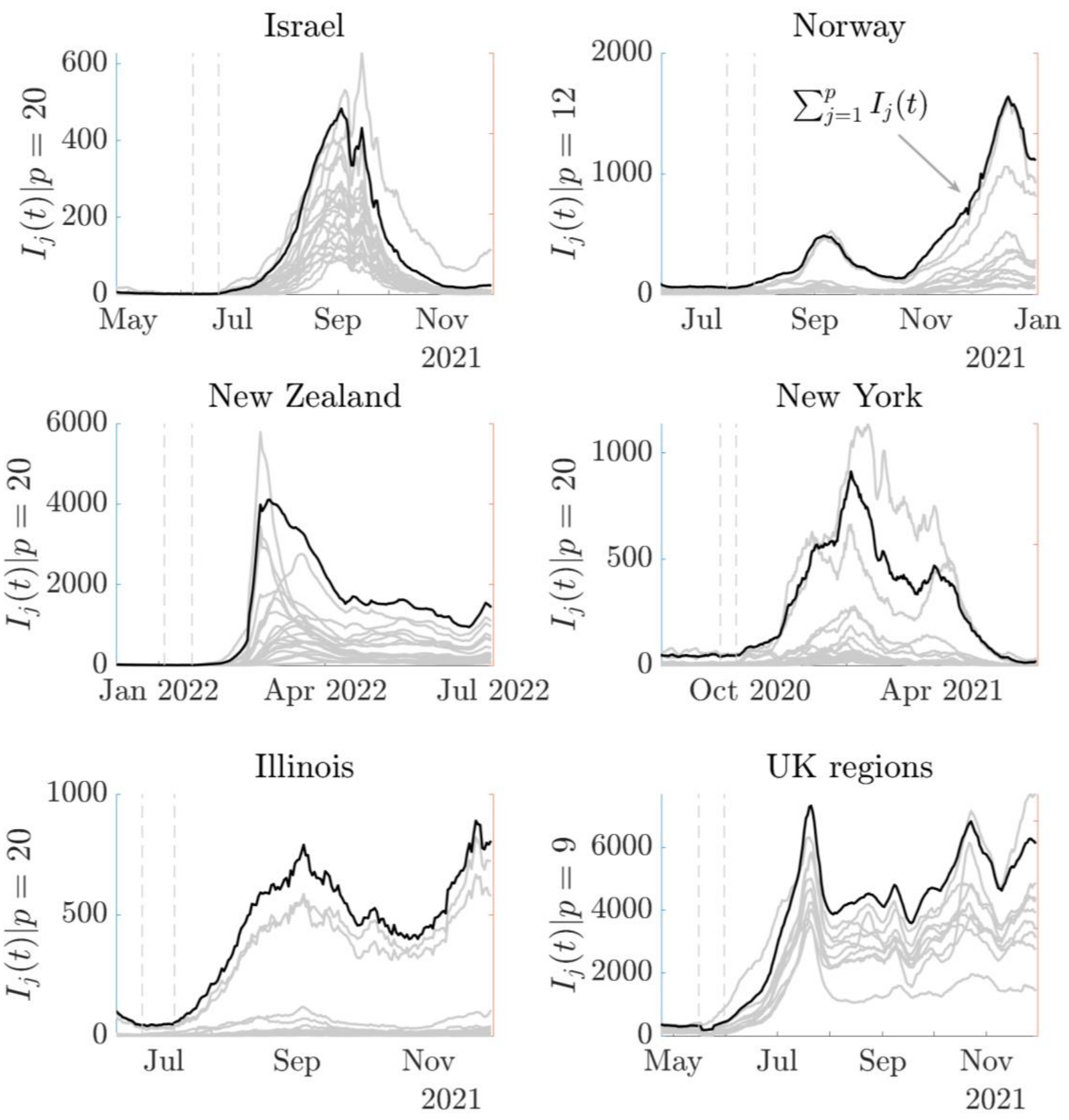


**Fig C: Incidence curves for 6 empirical COVID-19 datasets.** We plot new infections $I_j(t)$ over time $t$ for 6 case studies (data sourced from [8–13]) covering diverse countries, regions and states. Each dataset involves infections from $p$ groups, which we illustrate in grey. The black solid line, which is scaled down to be visible, shows the sum of infections from all groups. These curves underlie the results in **Figs 6-7** of the main text and in some scenarios show local infection increases that propagated into wider-scale resurgences. Vertical dashed lines show the periods over which we investigate resurgence signals in **Fig 7**.
